# Why COVID-19 models should incorporate the network of social interactions

**DOI:** 10.1101/2020.04.02.20050468

**Authors:** Helena A. Herrmann, Jean-Marc Schwartz

## Abstract

The global spread of Coronavirus Disease 2019 (COVID-19) is overwhelming many health-care systems. As a result, epidemiological models are being used to inform policy on how to effectively deal with this pandemic. The majority of existing models assume random diffusion but do not take into account differences in the amount of interactions between individuals, i.e. the underlying human interaction network, whose structure is known to be scale-free. Here, we demonstrate how this network of interactions can be used to predict the spread of the virus and to inform policy on the most successful mitigation and suppression strategies. Using stochastic simulations in a scale-free network, we show that the epidemic can propagate for a long time at a low level before the number of infected individuals suddenly increases markedly, and that this increase occurs shortly after the first hub is infected. We further demonstrate that mitigation strategies that target hubs are far more effective than strategies that randomly decrease the number of connections between individuals. Although applicable to infectious disease modelling in general, our results emphasize how network science can improve the predictive power of current COVID-19 epidemiological models.

## Introduction

The Coronavirus Disease 2019 (COVID-19), first identified in Wuhan, China in December 2019, has spread globally (Sun et al. 2020), overwhelming many national health care systems with an increasing number of serious and potentially life-threatening infections (Anderson et al. 2020). As a result, most governments have imposed mitigation and suppression strategies, such as social distancing and lock-down measures, to better control the spread of the virus. To ensure the effectiveness of these strategies, various epidemiological models are used to predict the spread of COVID-19 and to inform government policies (Ferguson et al., 2020; Hellewell et al., 2020; Sameni, 2020; Radulescu and Cavanagh, 2020; Simha et al. 2020; Zhao and Chen, 2020).

Most commonly, these models follow a general Susceptible-Infected-Removed (SIR) framework (Kermack and McKendrick, 1927; Bailey, 1975; Sameni, 2020). Infected individuals can only transmit the virus to susceptible individuals. Once infected individuals have recovered (or have passed away) they can no longer infect others and cannot be reinfected. With regards to COVID-19, the SIR model is often expanded to an SEIR model, which considers an additional Exposed (E) stage during which individuals have been infected but are not yet contagious (Radulescu and Cavanagh, 2020; Wu et al., 2020; He et al., 2020; Li et al., 2020). Many additional parameters, including clustering (Luo et al., 2020), age-heterogeneity (Chang et al., 2020; Radulescu and Cavanagh, 2020), changes in policy and control measures (Sameni, 2020; Zhao and Chen, 2020), and even meteorology (Jia et al., 2020) have been incorporated into the SIR and SEIR framework in an attempt to increase the predictive power of these models. Furthermore, there are models that consider an inherent randomness or stochasticity in the events that influence the model outcomes (Hellewell et al., 2020; Kucharski et al., 2020; Simha et al. 2020).

However, none of the above models, currently predicting the COVID-19 pandemic, take into consideration the structure underlying the human interaction network, thus limiting the possible mitigation strategies that can be considered (Manzo, 2020). Networks can be used to represent connections between individuals. A network of 10,000 nodes, for example, can be used to describe 10,000 individuals. The nodes in the network are connected by edges, which represent the interaction between those nodes. Two nodes sharing the same edge are considered to directly interact with one another and are referred to as neighbouring nodes. The number of edges of a node is referred to as the degree of that node (e.g. a node with two edges has a degree of 2). Nodes with many edges (and thus many neighbours) are referred to as “hubs”. It is critical to note that from a network analysis perspective, hubs do not necessarily refer to social gatherings. Hubs refer to individuals with many connections to other individuals; these connections do not necessarily occur at the same time. For example, a doctor who individually meets with 30 patients per day is considered a hub of 30 (i.e. a node of degree 30).

Current COVID-19 models are based on differential equations or random diffusions which assume that human interaction behaviours are generally homogeneous (i.e. alike). It is, however, well-documented that many biological interactions, including human face-to-face communications, are not homogeneous and not random (Barabási, 2009; Cattuto et al., 2010; Zhao and Bianconi, 2011; Zhang and Li, 2012; Manzo and Van de Rijt, 2020). In fact, the degree distribution of nodes (which captures differences in the number of connections between nodes) in a biological network can often be described by a power-law (Barabási and Albert, 1999). A power-law describes a relationship whereby one quantity varies as the power of another (e.g. the area of a square quadruples when the size of its sides is doubled). In networks, this property arises when there are few nodes with many edges and many nodes with few edges. Networks with this kind of property are known as scale-free. The rate of infection of COVID-19 has previously been shown to follow a power-law (Ziff and Ziff, 2020; Li et al., 2020); yet, so far scale-free networks have not been considered to model the disease.

When a disease spreads through a network, the infected nodes (I) can spread the disease only to their susceptible neighbours (S), who can then spread the disease to their susceptible neighbours, and so on. Using network science, Dezső and Barabási (2002) have previously identified that the best possible way to stop a virus from spreading in a scale-free network is to bias policies towards hubs (i.e. heavily connected nodes) in the network. They showed this using a Susceptible-Infected-Susceptible (SIS) model - a model that is arguably more suitable for the spread of computer viruses than infectious diseases. Here we re-assess the predictions of Dezső and Barabási (2002) using an SIR model, with the model parameters tailored specifically towards COVID-19. Our results demonstrate how mitigation strategies that directly target hubs in the network are far more effective than strategies that randomly decrease the number of connections between individuals. For example, reducing the total number of interactions that each individual in the network can have is potentially more effective than mitigating the number of interactions that an individual can have at the same time.

Although we have chosen model parameters that are based on the current COVID-19 pandemic, the model results cannot be considered a reliable prediction of the spread of this pandemic. Instead, they illustrate how network topology can improve the predictive power of such models. We propose that network topology should be combined with dynamic approaches in order to strengthen the predictive power of future pandemic models (Piccardi and Casagrandi, 2009). We further demonstrate how network topology can be used to suggest mitigation and containment strategies. In this sense, the results are generally applicable to a wide range of contagious infectious diseases.

## Results

We set up a scale-free network using the Barabási and Albert (1999) algorithm for a population of 10,000 (see Method and Materials for details).

The network is set up such that the majority of nodes in the network have less than *n* connections. Due to the scale-free structure of the network, there are some nodes that are heavily connected and have far more than *n* connections; these nodes are hereafter referred to as “hubs”. The value of *n* is an arbitrary cut-off and is dependent on the network structure. Here, we set *n*=8 so that hubs represent the 5-10% most connected nodes (Fig. 1).

**Figure 1:**
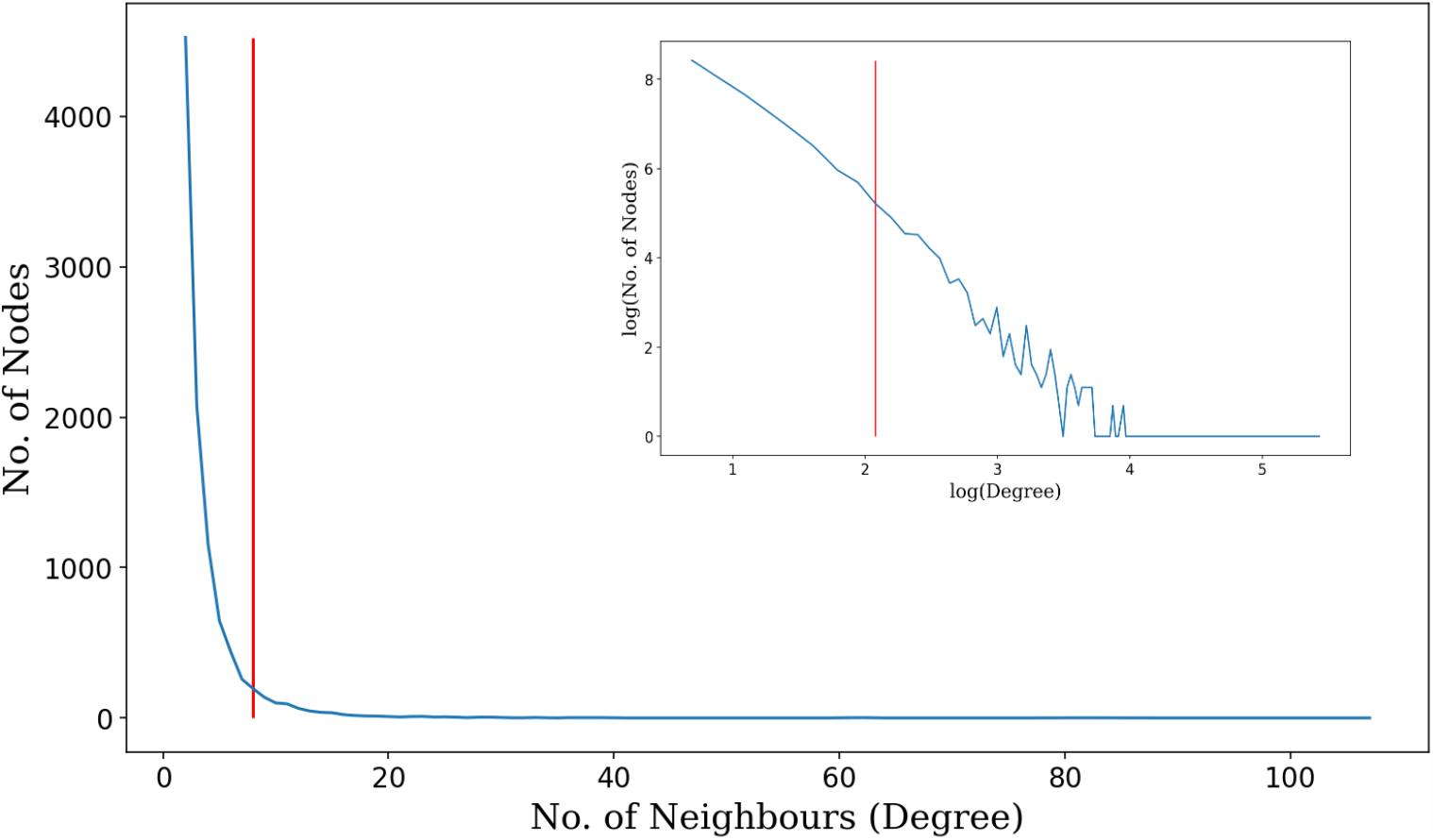
Example of degree distribution of a Barabási and Albert (1991) graph with 10 000 nodes generated using the *igraph* package (Version 0.8.0) in Python (Version 3.6.9) with a power of 1 and an edge per node connectivity of 2. The number of neighbours (degree) ranges from 1 to 107. This results in a power-law exponent of ∼2.46. Each individual has on average 4 neighbours. The same graph is shown on a logarithmic-logarithmic scale in the top right corner. Nodes with a degree greater than 8 (i.e. nodes with a degree to the right of the red cut-off) are considered hubs and make up less than 10% of the total number of nodes.

All nodes in the network were initially set to be in the susceptible (S) state. Randomly, the state of one node was changed to the infected (I) state, representing patient zero. Thereafter, at each time point, the infected node(s) can infect any of their neighbouring nodes, changing their state from S to I. This transmission can occur from 2-14 days after a node was infected (Lauer et al. 2020). The probability of transmission is highest from the 4^th^ to the 6^th^ day after being infected (He et al., 2020). After the 2-14 days of possible transmission, infected nodes (I) change to the removed (R) state. Nodes in the R state cannot become reinfected and can no longer infect others.

We then ran one set of models using the original scale-free network. We ran another set of models for which all hubs in the network had been contained; we randomly selected and retained 8 edges of each hub and removed all others. In this mitigated scenario, all nodes have a degree of 8 or less; we refer to this scenario as “Mitigation Hub”. To compare mitigation strategies that specifically target hubs to general mitigation strategies, we calculated the average number of edges that were removed in the mitigation strategy. We generated a third set of networks where we removed the same number of edges randomly within the scale-free network; we refer to this scenario as “Mitigation Random”. We ran the SIR model on all three network types (Scale-free, Mitigation Hub and Mitigation Random).

Figure 2 demonstrates how the number of susceptible, infected and removed nodes changes over time. There are two general outcomes in all three models: (i) the infection dies out quickly and does not spread throughout the network, or (ii) the number of removed nodes becomes high enough such that the spread of the infection is contained. In outcome (i) the majority of the population remains susceptible; in (ii) the majority of the population is infected and removed (i.e. has recovered or passed away). In outcome (ii) the population achieves a state known as “herd immunity” (Anderson and May, 1986; Kwok et al., 2020), whereby a high enough percentage of the population is immune to stop the virus from spreading. An illustrative example of the outcomes is provided, from a network point of view, in Figure 3.

**Figure 2:**
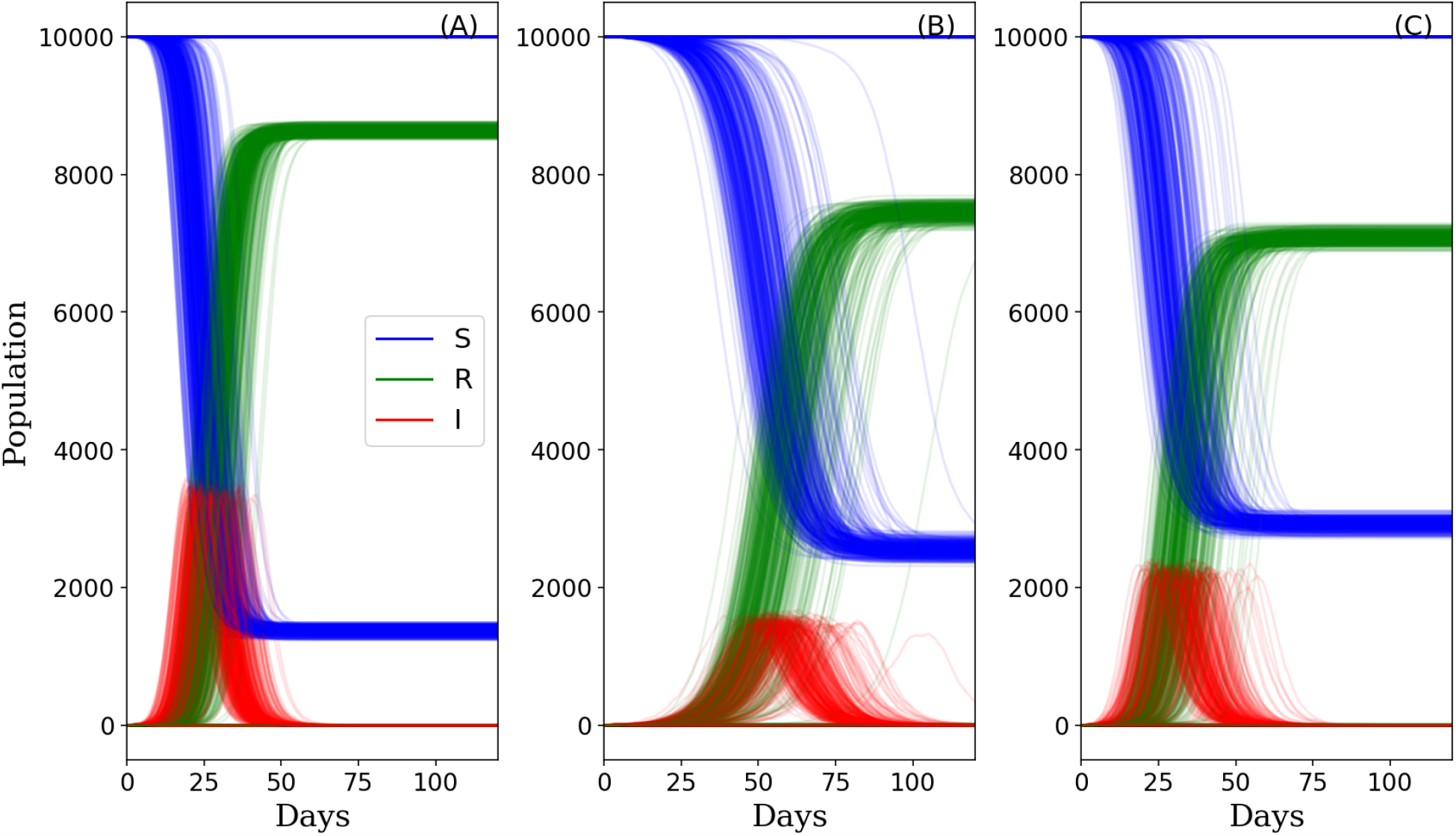
Population dynamics showing the number of infected (I, red), susceptible (S, blue) and removed (R, green) nodes in the network over time. The SIR model was run using: (A; Scale-free) the original scale-free network, (B; Mitigation Hub) the mitigated network where all nodes have 8 or less connections, and (C; Mitigation Random) the mitigated network where interactions were randomly removed. 500 model simulations were run for 120 days.

**Figure 3:**
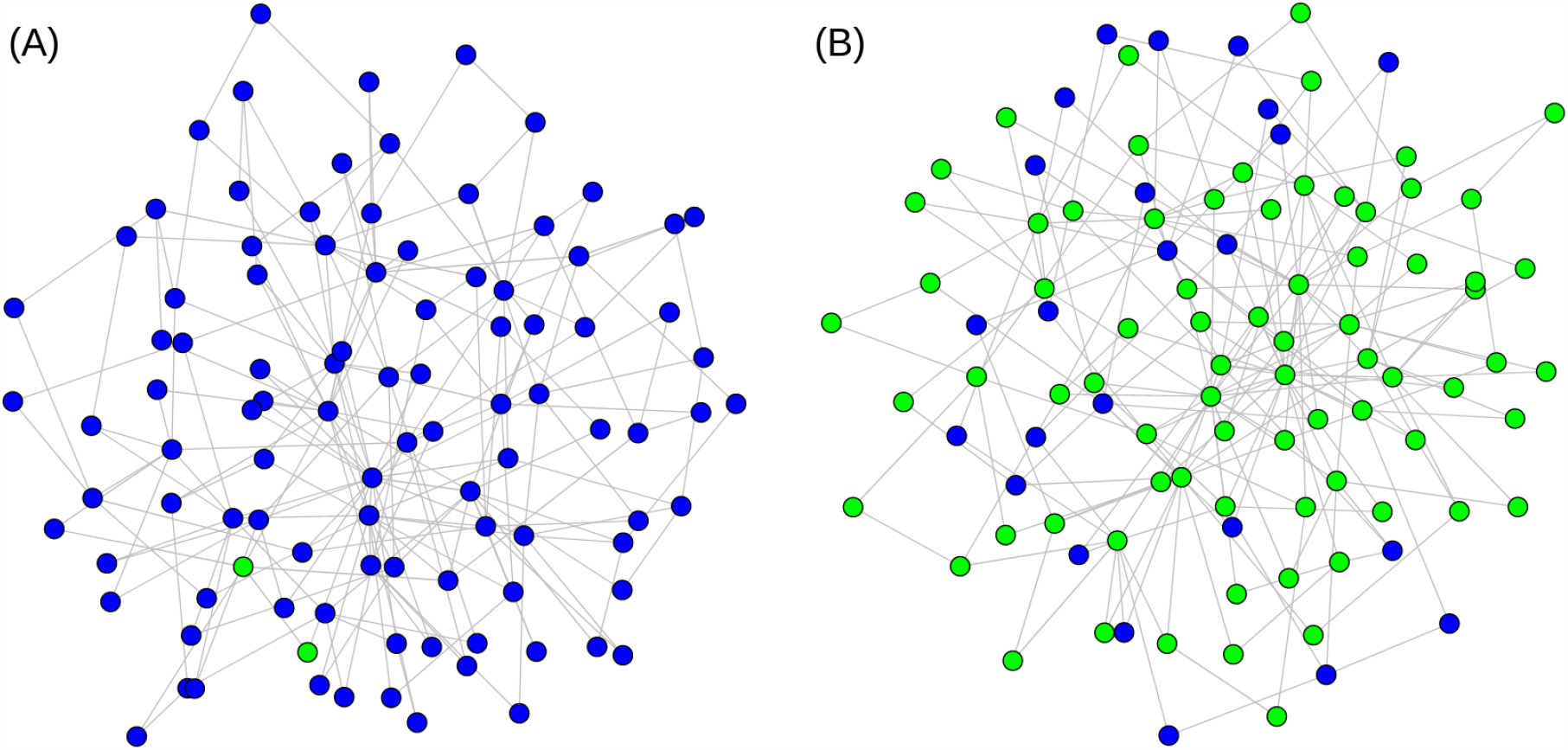
Illustrative examples of the two possible model outcomes. Removed (i.e. recovered or passed away) nodes are shown in green and susceptible nodes are shown in blue. For clarity we are showing a network with only 100 nodes. (A) No herd immunity is achieved because the spread of the virus ceases after infecting only two nodes. (B) Herd immunity is achieved and the spread of the virus ceases because the majority of nodes are removed and cannot be reinfected.

The distributions of the final state of the nodes in all model outcomes are shown in Figure 4A. The outcomes of the Mitigated Hub and Mitigated Random networks are similar in that they require a lower number of removed nodes to achieve herd immunity when compared to the outcomes of the scale-free network. However, the outcome of achieving herd immunity is also less likely when running the SIR model on the two mitigated networks, than on the original scale-free network (Fig. 4B).

**Figure 4:**
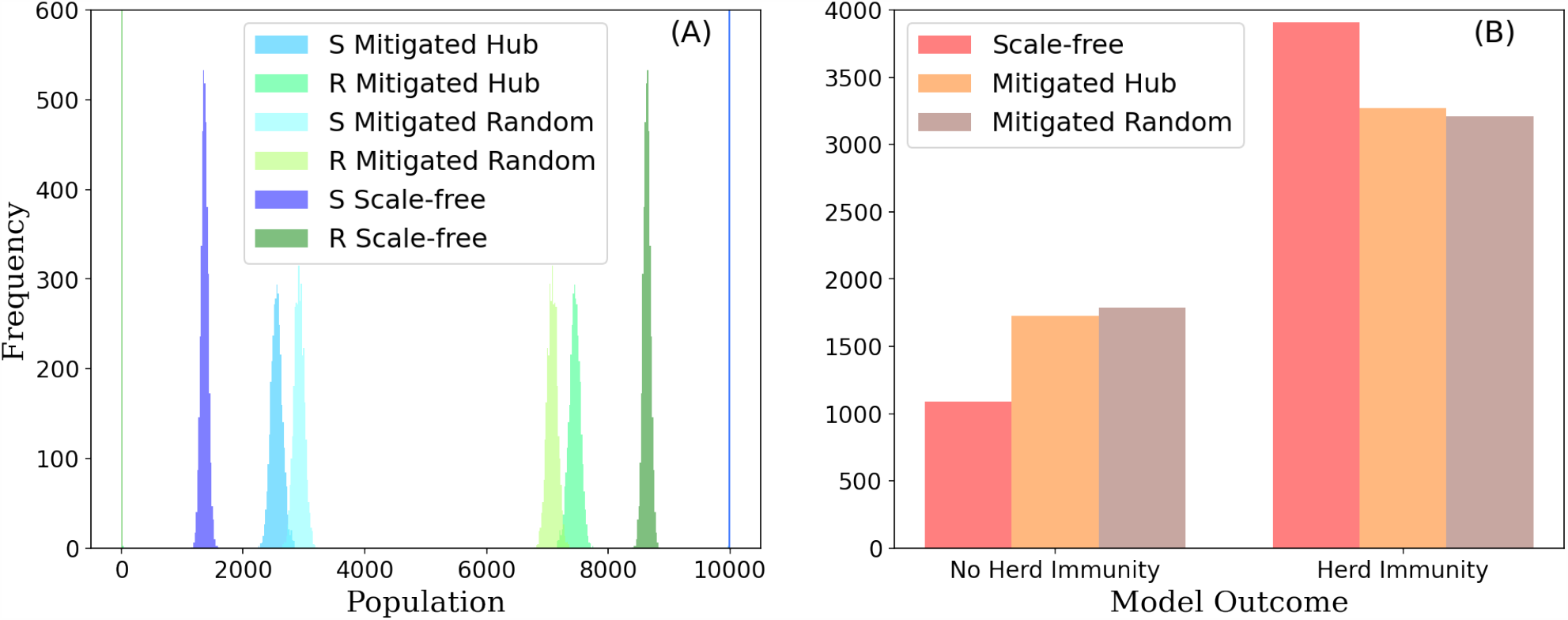
Bar charts of the possible model outcomes. (A) final states (R - removed; S - susceptible) of the model solutions of the three networks (Scale-free: the original network; Mitigated Hub: where edges were removed from the scale-free network such that all nodes have no more than 8 connections, Mitigated Random: where the same number of edges were removed randomly). (B) final states of the two model outcomes (No Herd Immunity - where the infection dies out early, before spreading, and the majority of the population remains susceptible; Herd Immunity - where the infection stops spreading because the majority of the population is immune). 5,000 model simulations were run for 170 days.

When running the SIR model on the scale-free network, over 80% of the population needs to become infected for herd immunity to be achieved (Fig. 2A). In the mitigated scenarios herd immunity can be achieved with around 70-75% of the population becoming infected (Fig. 2B,C). This is because the number of connections between nodes is limited. When specifically limiting the connections of hubs, the rate at which the virus spreads through the network is slowed down (Fig. 2B & Fig. 5). When removing hubs from the network, the onset of more than 1% of the population being infected occurs later than with any of the other networks (Fig. 5A). Also, in the Mitigated Hub scenario, the rate of infection is slower such that, at the peak of infection, a significantly lower percentage of nodes is infected with the virus (*p* < 0.01, Kruskal-Wallis, n > 3320) (Fig. 5B). This means that when the SIR model is run on the Mitigated Hub network, the time for which more than 1% of the population is infected is longer and the peak of infection occurs later in time (Fig. 6); thus, the curve is flatter (Fig. 2B).

**Figure 5:**
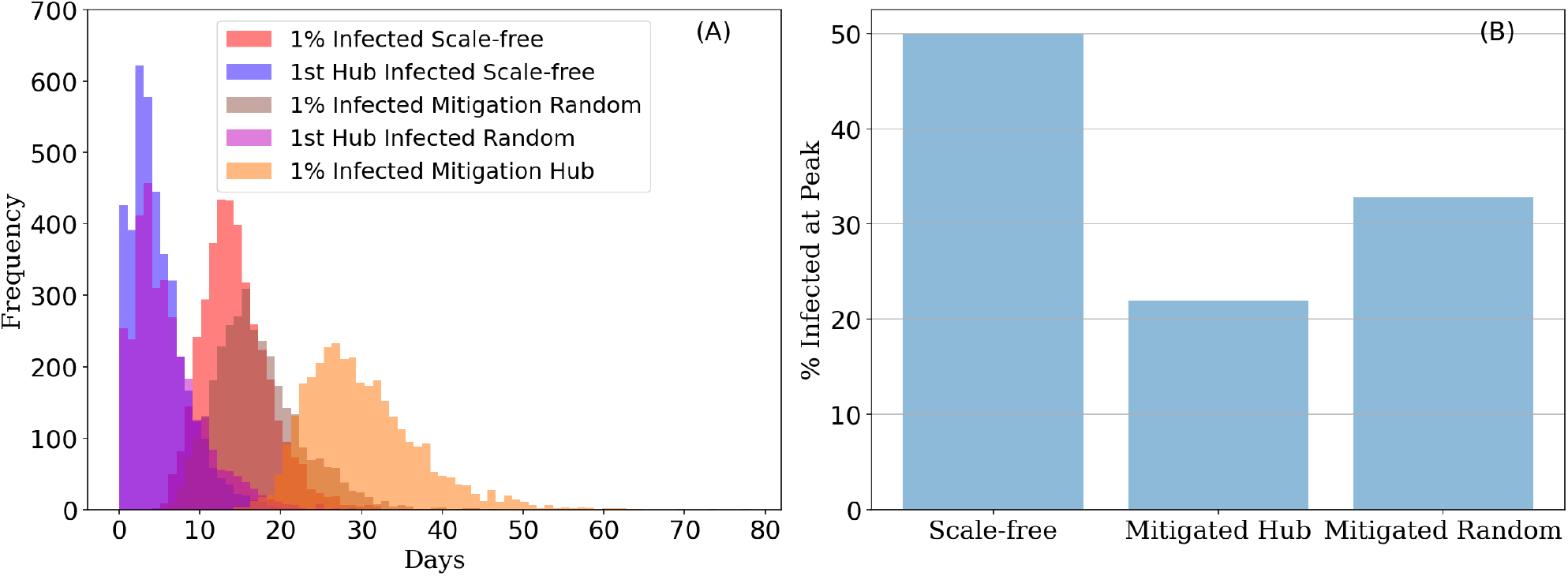
Time at which more than 1% of the population is infected in the Herd Immunity outcome. The SIR model was run on the three networks (Scale-free: the original network; Mitigated Hub: where edges were removed from the scale-free network such that all nodes have no more than 8 connections; Mitigated Random: where the same number of edges were removed randomly). (A) frequency distributions of the onset of time after which more than 1% of the population is infected for the three networks (Scale-free - red; Mitigation Hub - orange; Mitigation Random - brown) and of the time at which the first hub (i.e. the first node with more than 8 connections) is infected in the scale-free network (blue) and in the Mitigation Random network (purple). (B) the number of nodes that are infected at the peak of the infection divided by the final total number of removed nodes, shown as a percentage. For both (A & B), 5,000 model simulations were run for 170 days.

**Figure 6:**
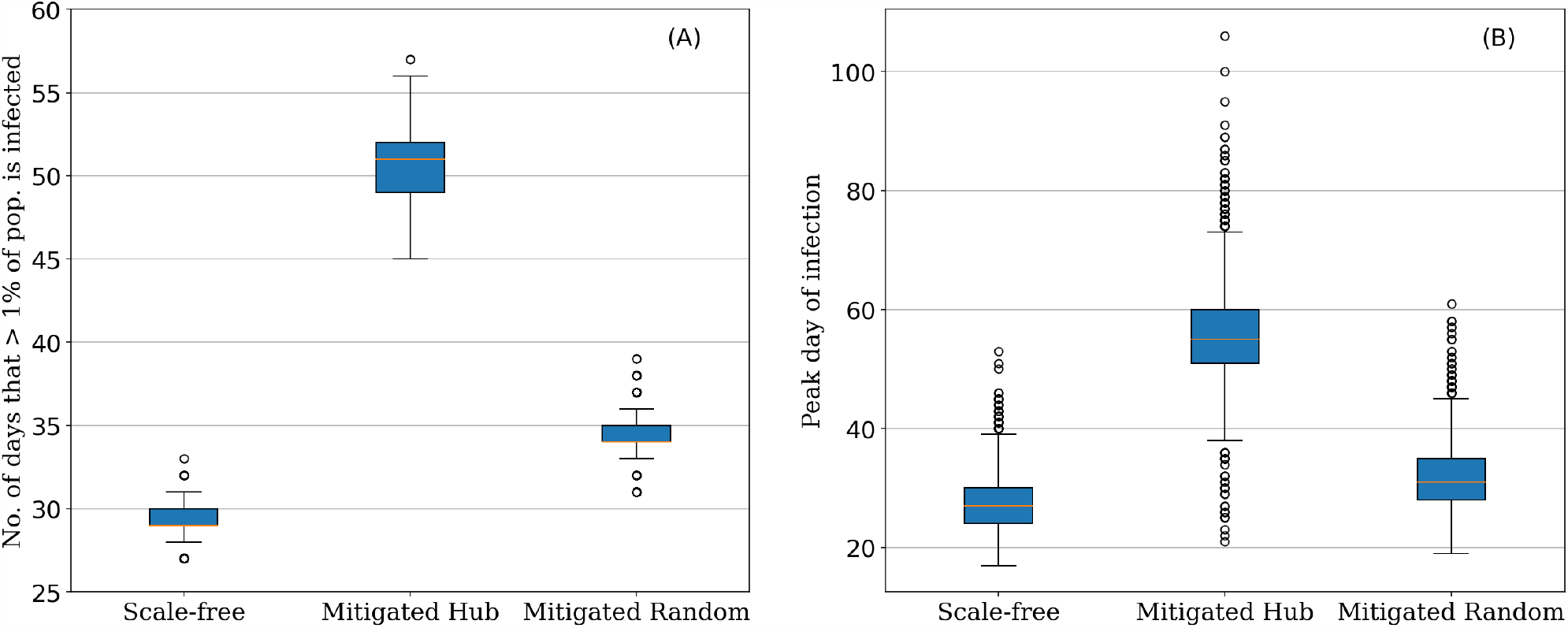
Duration of peak infection and the day at which the greatest number of nodes are infected when running the SIR model on three different networks (Scale-free: the original network; Mitigated Hub: where edges were removed from the scale-free network such that all nodes have no more than 8 connections; Mitigated Random: where the same number of edges were removed randomly). (A) the average total number of days for which more than 1% of the population is infected. (B) the average number of days after onset of the infection when the greatest number of nodes is infected at the same time. Both (A & B) were calculated from 5,000 model runs; error bars show the respective standard deviation. Only outcomes where herd immunity was achieved were considered.

The onset of more than 1% of the population being infected occurs just after the first hub has been infected (Fig. 5A). For the Mitigated Hub network (which contains no hubs), the time of onset at which more than 1% of the population becomes infected is more varied, as indicated by the wider distribution shown in orange.

All our networks consist of a population of 10,000 individuals, thus representing a small community. Including more individuals into the networks would not change the proportionality of our results. However, previous network analyses have suggested that networks representing human interactions across wider geographical regions should consider transitivity (Serrano and Boguna, 2006; Friemel, 2011). Transitivity is a measure of the density of connections in a network (Wasserman and Faust, 1994) and is often also referred to as the clustering coefficient (Newman, 2018). The higher the transitivity of a network, the denser are its connections. We calculated the transitivity for all our generated networks and found that when network transitivity is high, the peak of infection rises and ceases sooner (Fig. 7). Networks with no hubs (Mitigation Hub) are the least densely connected network and thus the peak of infection is flatter (Fig. 2 & 7).

**Figure 7:**
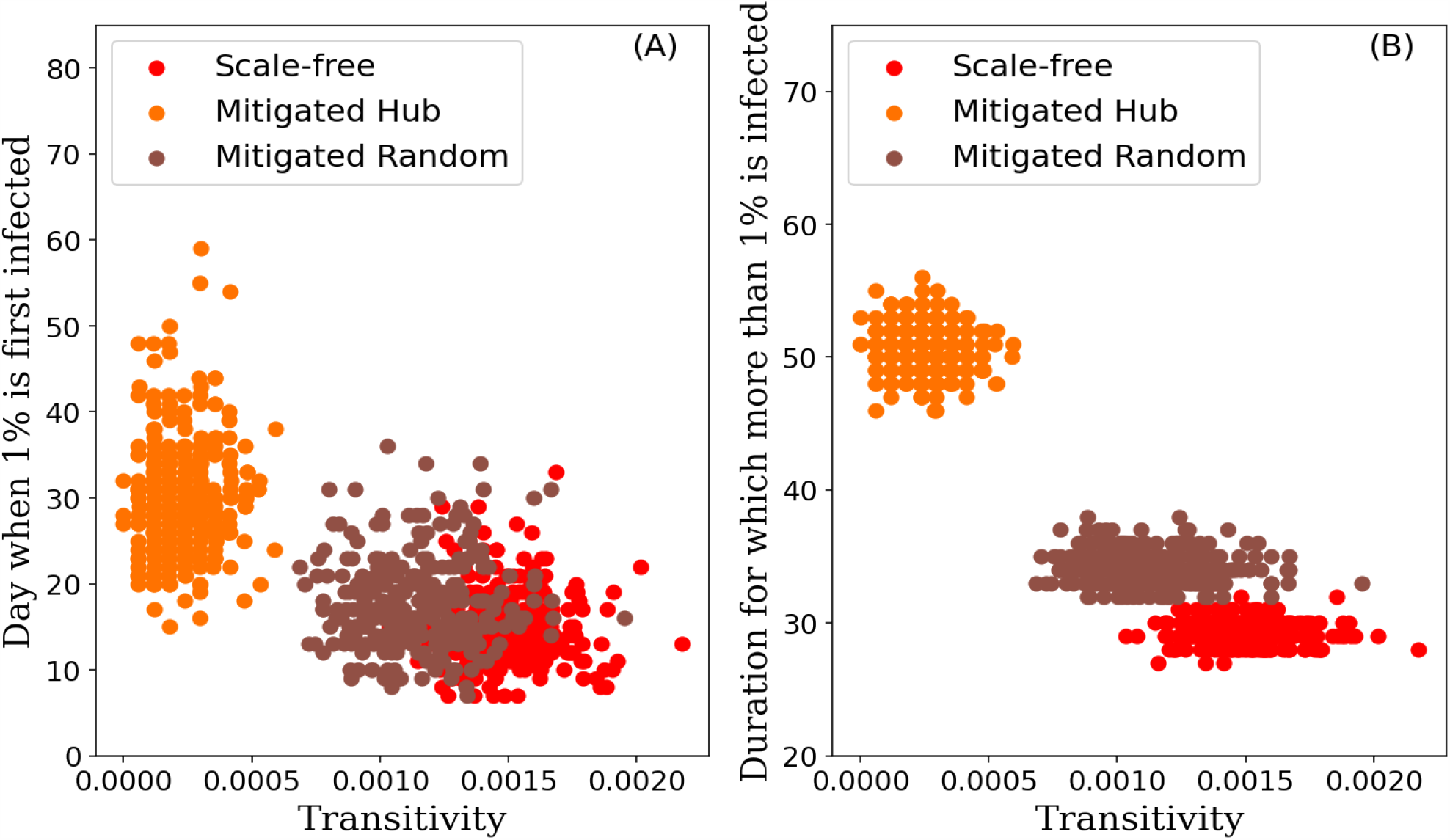
The effect of network transitivity (i.e. clustering) on the spread of an infection. Transitivity of the generated networks (Scale-free in red - the original network; Mitigated Hub in orange - where hubs with more than 8 connections have been removed; Mitigated Random in brown - where edges have been removed randomly) plotted against (A) the time (day) when more than 1% of the population is first infected in the herd immunity outcome and (B) against the total time (days) for which more than 1% of the population is infected in the herd immunity outcome. 300 model simulations were run for 170 days.

## Discussion

We have shown that incorporating a scale-free network structure of human interactions into an SIR model of COVID-19 can provide novel insights into the potential strategies and policies for mitigating and suppressing the spread of the virus. Our results demonstrate that targeting hubs in a network has the potential to slow down the rate of infection and to “flatten the curve”. Removing hubs from a scale-free network markedly reduces the number of infected individuals at a time and could therefore reduce the strain on health care systems, if implemented early on. We show that removing edges from hubs rather than from random locations in the network is a more successful strategy for slowing down the spread of the virus. Thus, limiting the total number of interactions that each individual in a network can have, could be an effective policy when trying to reduce the number of infections at a time. Our results are in agreement with those by Monzo and Van de Rijt (2020) who constructed an SEIR model based on a specific social interaction network based on a survey conducted on over 2000 individuals in France.

We further demonstrate that the onset of the peak of infection occurs shortly after the first hub is infected. If, by chance, no hub is infected early on, then the infection dies out or the peak is noticeably delayed until a hub is infected. These results align with data coming from Italy, which suggest that the COVID-19 epidemic must have entered the region far earlier than when the first case was officially detected (Cereda et al., 2020). According to their study, the virus could have circulated for several weeks with the number of infected people remaining low, until it started to rise sharply.

Our results further align with those obtained by Dezső and Barabási (2002) who modelled the spread of a virus using an SIS model and suggested targeting hubs as a primary mitigation strategy. Their model, however, does not provide the option of a herd immunity outcome as it does not consider a removed state. At the time of writing, the extent to which individuals can become re-infected is still an open discussion. It is assumed that some level of immunity exists in individuals that survive an infection (Bao et al., 2020; Kirkcaldy et al., 2020, Zhao et al., 2020). All of our SIR models present two possible outcomes: either the infection dies out quickly and the majority of the population remains susceptible, or the majority of the population gets infected and herd immunity forces the virus to stop spreading. If vaccines are available, herd immunity can be achieved quicker. Vaccines would allow nodes in the network to move from a susceptible straight into a removed (i.e. recovered) state. Unfortunately, no vaccine for COVID-19 is available at the time of writing (Amanat and Krammer, 2020; Pompetchara et al., 2020). If future research shows that it is possible to get re-infected with the disease, then it may be necessary to adapt the model to allow individuals that survived the disease to move back into a susceptible stage after a specified time period.

We did not opt for an SEIR model, but instead set the probability of infection very low for the first few days after contracting the virus. This matches the current available data on COVID-19 (He et al., 2020) and has a similar effect as an SEIR model would have. Furthermore, we did not distinguish between recovered and passed away in the models. From a modelling perspective, whether a node recovers or dies has the same implication: it cannot get reinfected nor can it spread the virus. From a human health perspective, the fact that the number of removed nodes is likely to include a number of deaths (depending on the fatality of the virus), paints a potentially sombre picture for the model outcomes in which herd immunity is achieved. For COVID-19, the current case fatality rates of Germany and Italy were calculated to be 0.2% and 7.7%, respectively (Lazzerini and Putoto, 2020). Evidently, these figures are dependent on the number of cases tested as well as other factors. Anyway, the potential magnitude of fatality rates implied by these numbers does suggest that mitigation strategies that lower the number of infected people required for herd immunity may be necessary to keep high-risk citizens safe from the infection. In this case, a combination of hub specific and equally applied mitigation (i.e. random) strategies should be considered. Hub specific mitigation strategies are more likely to lower the peak of infection, whereas equally applied mitigation strategies are more likely to increase the number of individuals that never contract the disease.

Evidently, the results of our model come with a set of assumptions that are not necessarily valid under the current and evolving circumstances of the spread of COVID-19. For example, our network analysis considers the number of connections to be constant across time. This however is unlikely, given that even without government enforced policies, individuals are likely to reduce their face-to-face interactions once they fall ill. Workers providing essential services, like healthcare and delivery personnel, on the other hand, may increase their number of interactions with the spread of the virus. This could result in hubs becoming even more connected over time. Furthermore, both our discussed mitigation strategies are applied before the onset of the infection, in order to illustrate a point. A more pragmatic model should consider how network dynamics and policies enforced during the pandemic can alter the degree distribution of the network over time (Piccardi and Casagrandi, 2009; Barrat et al. 2013).

We showed that the transitivity in a network (i.e. the density of its connections) correlates with the sharpness of the infection peak. The onset of the peak occurs later and is slowed down in networks where hubs have been removed, because these networks have a reduced transitivity. However, our results did not consider modularity, also known as community structure. Previous research has shown that a community structure in networks can reduce the risk of a pandemic (Eguiluz and Klemm, 2002; Huang and Li, 2007; Stegehuis et al., 2016). We therefore recommend that, when upscaling our approach to model the spread of an infection across a wider geographical network, community structures are implemented.

In conclusion, we here present a fundamental example for why the network structure that underlies human interactions should be taken into consideration when modelling the spread of a virus, such COVID-19. We emphasize that the model results should not be used as predictions of the spread of COVID-19 but as a guide on how current epidemiological models could be improved. Incorporating network science with the current dynamic models of COVID-19 is likely to improve their predictive power. Doing so will allow for better-informed suggestions for disease mitigation and suppression, such as reducing the number of hubs in a network. Strategies that consider the underlying network structure of human interactions will allow the implementation of more tailored policies, attempting to effectively deal with COVID-19 and other pandemics.

## Materials and Methods

### Network Generation

Barabási and Albert (1999) graphs of 10,000 nodes were generated using the *igraph* (Version 0.8.0) package in Python (Version 3.6.9) with a power of 1 and an edge per node connectivity of 2. These are scale-free networks and the node degree distribution follows a power law (e.g. Fig 1). We then generated a set of networks where we removed all hubs from the original scale-free versions. If a node in the scale-free network contained more than 8 edges, we randomly removed edges from that node, such that it only had 8 edges left in total. We refer to these networks as “Mitigated Hub” networks. When generating 5,000 Mitigated Hub networks, we removed, on average, 4613 edges per network. As a control we also generated a set of networks where we removed that same amount of edges randomly (i.e. not targeting hubs). We refer to these networks as “Mitigated Random” networks. In total we generated 3 sets of networks (Scale-free, Mitigated Hub, and Mitigated Random). We generated 5,000 networks of each set. Network properties, such as transitivity, were also calculated using the *igraph* package. Network transitivity, also known as the global clustering coefficient, (C) was calculated such that C = tr(A^3^) / ∑_i≠j_ (A^2^)_i,j_ (Newman, 2010).

### SIR Model

The nodes of the networks can be either in a susceptible (S), an infected (I) or a removed (R) state. Initially, all nodes were set to be in state S. Then at the first time point, we randomly choose one of the nodes in the network to be in the infected state. This node represents patient zero who first contracts the virus from a non-human source. We then update the model such that at each iterating time point infected nodes can infect their susceptible neighbouring nodes with a given probability *p*. Infected nodes can remain infected for 2 to 14 days (Lauer et al. 2020). The length of time that a node is infected is randomly chosen, with equal probability. Once the node is no longer infectious it is set to be in the R state and cannot be infected again. The probability with which each infected node can spread the virus to its susceptible neighbours peaks at around 5 days after contracting the infection. To match the current available data for COVID-19 (He et al., 2020; Li et al., 2020), we set *prob* = [0.01, 0.01, 0.1, 0.2, 0.3, 0.3, 0.3, 0.25, 0.2, 0.15, 0.1, 0.05, 0.01, 0.01], representing the probability of infection on each of the 14 days, respectively. All models are run to completion (i.e. until the infection dies out). Once the infection dies out, all nodes are either in the R or S state. We ran the here described SIR model on each of the generated networks (see Network Generation). We used the Kruskal-Wallis test, as implemented in the *scipy* package (Version 1.4.1) to test for significant differences between distributions.

## Data Availability

The code used to generate all the data presented in this study is available on Github (https://github.com/HAHerrmann/NetworkEpidemics) and Zenodo (DOI: 10.5281/zenodo.3736466).

## Data Availability

The code used to generate all of the data presented in this study is available on GitHub (https://github.com/HAHerrmann/NetworkEpidemics) and Zenodo (DOI:10.5281/zenodo.3736466).

DOI:10.5281/zenodo.3736466

## Acknowledgements

HAH is supported by a Biotechnology and Biological Sciences Research Council (BBSRC) Doctoral Training Partnership stipend (BB/M011208/1). We thank Alexander L. Herrmann for valuable discussions on the topic.

## Author Contributions

JMS conceived the idea. JMS and HAH conducted the analyses. HAH wrote the manuscript. Both authors reviewed the manuscript.

